# The Effect of Negative Pressure on IOP in the Living Human Eye

**DOI:** 10.1101/2022.06.27.22276880

**Authors:** Massimo A. Fazio, Gianfranco Bianco, Udayakumar Karuppanan, Meredith Hubbard, Luigi Bruno, Christopher A. Girkin

**Affiliations:** Department of Ophthalmology and Visual Sciences, The University of Alabama at Birmingham, School of Medicine, Birmingham, AL, USA; Hamilton Glaucoma Center, Shiley Eye Institute, Viterbi Family Department of Ophthalmology, University of California San Diego, La Jolla, CA, USA; Department of Neurobiology, The University of Alabama at Birmingham, School of Medicine, Birmingham, AL, USA; Department of Mechanical, Energy, and Management Engineering, The University of Calabria, Arcavacata di Rende, Italy

**Author notes:** **Correspondence** Massimo A. Fazio, Ph.D., Department of, Ophthalmology and Visual, Sciences, 1720 University Blvd, Birmingham, AL, United States, 35294.

## Abstract

**Purpose:** 

To quantify the effect of negative pressure applied to the anterior surface of the eye on absolute IOP.

**Subjects:** Participants, and/or Controls. Three eyes from three research-consented brain-dead organ donors.

**Methods:** Air-tight goggles connected to a negative pressure (NP) pump (Mercury Multi-pressure Dial (MPD); Equinox Ophthalmic, Inc., CA) were tested on three research consented brain-dead organ donors. The MPD was set to generate a vacuum of -20mmHg. A baseline IOP of 10, 20, and 30mmHg was sequentially set by adjusting the height of a balanced salt solution bottle connected to the cornea through a 20G needle. IOP was manually annotated at time = 0s, right before turning on the vacuum pump; after a few seconds with the vacuum pump ON; at 60s; at 120s, right before turning the pump OFF; at 240s with the pump OFF (recovery). Three repetitions per each test at varying baseline IOP were taken for a total of nine series of measures per eye.

**Main Outcome Measures:** IOP change with exposure to NP.

**Results:** Eye1 and 3 showed an immediate reduction in IOP at all baselines immediately following activation of the MPD NP pump; Eye2 showed an opposite response to NP. Eye1 and 3 showed a reduction in IOP at all baselines with NP pump ON for 60s and 120s, while Eye 2 showed a consistent increase in IOP. After 120s from turning NP OFF (time=240s), IOP partially recovered to its baseline in all eyes.

**Conclusions:** This study is the first to evaluate changes in manometrically-measured intracameral IOP due to NP applied to the ocular surface in living conditions. The inconsistent response of IOP following exposure to negative pressure warrants further investigations on the mechanism underlaying IOP lowering by NP.

## Introduction

Intraocular pressure (IOP) is a major risk factor for the development and progression of glaucoma and reduction in IOP remains the only proven treatment to slow or halt the disease.^1, 2^ While other factors are involved^3-5^, even in patients who develop or progress with glaucoma with normal pressure, lowering of IOP is beneficial. IOP provides the loading force to the ocular coat (sclera, cornea, and optic nerve head (ONH)) needed to maintain the consistent shape of the globe required for pristine optics and is counterbalanced posteriorly by cerebrospinal fluid pressure^6-8^ (CSFP) in the region of ONH and orbital pressure, and anteriorly, by atmospheric pressure.

Associations between changes in atmospheric and orbital pressure with IOP, and the forces IOP imparts to the retina, vasculature and ONH have been observed in aviators^9^ and divers^10^ for over fifty years.^9, 11-13^ Atmospheric and orbital pressures represent additional mechanical forces,^14-16^ along with IOP and CSFP, that impact all ocular tissues. While changes in atmospheric pressure occur mostly in specialized conditions (e.g., diving, altitude changes), alteration in atmospheric pressure around the eye has been proposed as a novel treatment of glaucoma to non-invasively reduce IOP.^17-19^

Based on this potential impact of atmospheric pressure on IOP, interest has developed in evaluating the application of goggles that apply negative pressure to the ocular surface simulating a drop in atmospheric pressure to provide a potential non-invasive therapeutic option to reduce IOP in glaucoma^16-22^. Computational studies have suggested that this approach should lower IOP due to scleral expansion acutely and decrease episcleral pressure over longer periods.^16^ Clinical evaluation of an investigational device, has found a reduction in IOP measured with pneumotonometry across a latex membrane imbedded within the goggle system.^20, 21^ The current study measured manometric IOP directly in the anterior chamber while applying negative pressure using the MPD to determine the acute impact of negative periocular pressure on IOP in three research-consented brain-dead organ donors (BDOD) immediately prior to organ recovery.

## Materials And Methods

The study was developed within the Legacy of Hope organ recovery center at the University of Alabama at Birmingham. All components of this study adhered to the Declaration of Helsinki and were approved by the UAB Institutional Review Board and Legacy of Hope Research Review Board.

This unique center was developed as a centralized site for organ procurement and allows for intensive care level management of organ donors maintained on life support to improve the outcome of organ donation. While agonal effects in the eyes are present in many organ donors, many braindead organ donors have completely normal eye exams with normal retinas on fundoscopic examination, optical coherence tomography (OCT) and OCT angiography (OCTA).

Legacy of Hope consents organ donors for research on tissues and organs and runs several research protocols, including the recent xenotransplant into the first cadaveric donor model.^23^ We have developed the Living Eye Project within this center as a platform to test the living human eye in-vivo before organ procurements ^12^.

For this study, we utilized the MPD device modified to allow for anterior chamber cannulation for the measurement and adjustment of IOP (Fig. 1). Three research consented organ donors were utilized in this study. The eyes in these donors were not eligible for transplant. Following clearance from Advancing Sight, Inc., formerly the Alabama Eye Bank, to ensure donor eligibility, the anterior chamber was cannulated using a 20-gauge anterior chamber maintainer (Anodyne Surgical, Inc.; O’Fallon, Missouri) affixed to pass through the flexible skirt of the modified MPD goggles and inserted into the eye through a clear corneal incision made with a 1mm keratome. This anterior chamber maintainer is designed for a water-tight seal using this keratome. Once in place, the wound was checked under microscopic visualization for any leakage. The goggles were then place on the orbital rim to ensure an airtight seal which was verified by the device. The infusion line was connected to an inline pressure gauge (X2Pi, Crystal Engineering; San Luis Obispo, CA) and connected to a bottle of balanced salt solution (BSS) that could be positioned at varying heights so that IOP could be varied by adjusting the fluid column height. IOP was manually annotated by taking note of the pressure gauge reading at specific times during testing (described below).

**Figure 1.**
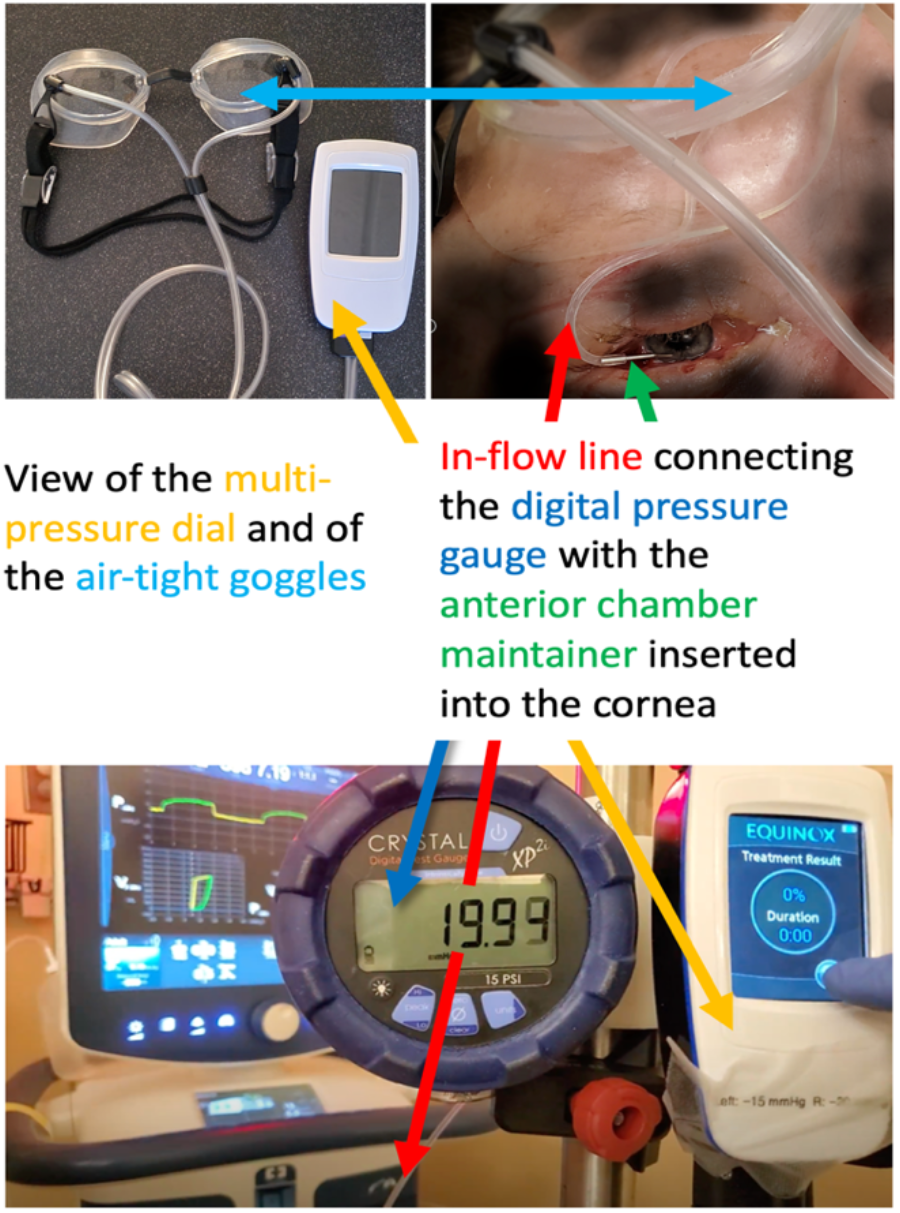
Layout of the testing set up. **Top left**: view of the multi-pressure dial (MPD) and air-tight goggles. **Top right**: view of infusion line tube passing through air-tight goggles and inserting via an anterior chamber maintainer inside the anterior chamber of the donor’s eye. **Bottom**: view of the pressure gauge manometer for measuring intraocular pressure, and of the MPD negative pressure pump.

In each donor, IOP was initially set at 10mmHg by adjusting the BSS bottle height appropriately by using a laser light leveler. Once baseline IOP was stable at approximately 10mmHg, the vacuum pump connected to the MPD goggles was activated to apply a nominal vacuum pressure of -20mmHg from atmospheric; this negative pressure was applied for 2 minutes counted by a portable stopwatch. After the 2 minutes with the MPD vacuum pump activated, the pump was turned off and IOP was monitored for another 2 minutes. IOP, as read at the pressure gauge display, was recorded at five specific points in time (stages) : 1) at baseline IOP just before activation of the vacuum pump; 2) immediately following (∼1 second (s)) activation of the MPD vacuum pump; 3) after one minute (60s) and 4) after two minutes (120s) following activation of the vacuum pump; and 5) after two minutes following turning off the MPD vacuum pump (240s). This sequence of vacuum pressure changes and IOP readings was repeated three times. This repeated sequence was performed at baseline IOPs of 10, 20, and 30 mmHg. Hence, a total of nine measures per eyes were collected.

All the procedures were recorded by a portable video camera capturing readings of the pressure gauge and MPD displays, as shown in Fig. 1.

To ensure that IOP changes seen with negative pressure were not due to expansion of the infusion line within the negative pressure space of the MPD goggles, in the first tested eye (Eye1), after the above testing protocol, the following procedure was performed: the anterior chamber catheter was removed from the eye; the infusion tubing was completely occluded and placed into the goggle chamber; the testing protocol was then repeated at 10, 20 and 30 mmHg with the occluded tube outside of the eye but within the negative pressure space; IOP was annotated at each repetition (results later shown).

### Statistical analysis

The change in manometric intracameral IOP with negative pressure against its baseline value was computed by a linear mixed-effect model (LME) with a random intercept per study subject. Statistical analyses were performed in R (R Foundation for Statistical Computing: Vienna, Austria; lme4 package). A p-value of < 0.05 was considered significant.

## Results

The test protocol defined in the method section was applied to three donors. Age, sex, race, and BMI data are reported in Table 1. The measured IOP for the three eyes, three baselines, and three repetitions are shown in Table 2. Figure 1 illustrates the effect of negative pressure (−20mmHg) over time on IOP for each of the tested eyes and at varying baseline IOPs with each plot overlayed on each other the three repetitions of IOP measurements. Noticeably, the three repeated measures were highly repeatable with each donor and across all the baseline IOPs.

**Table 1.** Demographics of the three research-consented brain-dead organ donor (BDOD) eye

| BDOD ID | Age range | Sex | Race | BMI |
| --- | --- | --- | --- | --- |
| Eye 1 | 30-40 | M | ED | 31 |
| Eye 2 | 50-60 | F | ED | 52 |
| Eye 3 | 40-50 | F | AD | 27 |

**BDOD ID:** brain-dead organ donor identifier; **BMI:** body mass index; **ED:** European descent; **AD:** African descent

Eye 1 and 3 (Fig. 2, 1^st^, and 3^rd^ columns) showed an immediate reduction in IOP at all baselines following activation (time= 0s) of the device applying negative pressure (shown in Fig. 1). Eye 2 (Fig. 2, 2^nd^ column) showed an inconsistent response with a slight initial decrease in IOP at a baseline IOP of 10mmHg and an increase in IOP at baseline IOPs of 20 and 30mmHg.

**Figure 2.**
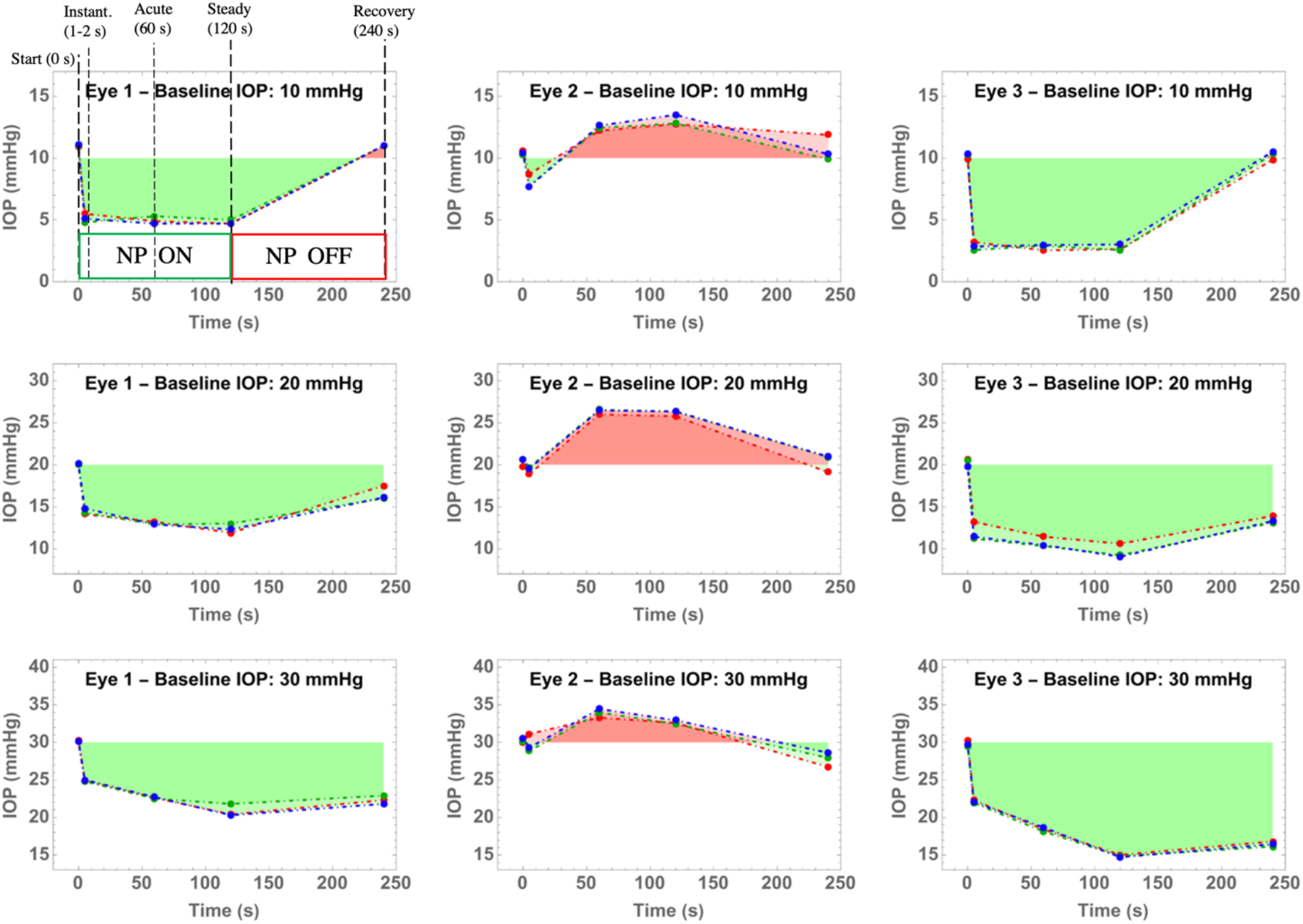
Change in IOP as it changes with negative pressure and baseline IOP (10, 20, 30mmHg) in the three eyes tested in living conditions (grouped by column). IOP was discretely measured at five stages (time points: 0s, 1-2s, 60s, 120s, 240s). The NP pump was ON and set at -20mmHg for the first 120 seconds, then it was turned OFF for the 120 to 240 seconds interval. Areas of the plots highlighted in **green** indicate that the IOP punctual value is lower than its respective baseline; highlighted in **red** indicates higher IOP than baseline. The five testing stages are shown as overlay only on the top-left plot. **IOP**: Intraocular pressure; **NP**: negative pressure; **mmHg**: millimeter of mercury; **s**: seconds.

After one minute of negative pressure, Eye 1 and 3 showed a reduction in IOP at all baseline IOP levels after 60 seconds from the application of negative pressure, while Eye 2 showed a consistent increase in IOP across all baseline pressures.

The IOP response at two minutes showed steady and similar reductions compared to the IOP at 1 minute following exposure to negative pressure in all eyes. Follow removal of negative pressure, Eyes 1 and 3 showed full recovery to baseline values at baseline 10mmHg at 240 seconds. Partial recovery was observed for baseline in these two eyes for at baseline pressure of 20mmHg, with a larger sustained reduction observed in these two eyes at a baseline IOP of 30mmHg.

In the eye with pressure elevation following the application of negative pressure, pressure returned to baseline IOP at 10 and 20mmHg and was slightly lower than baseline at 30mmHg. No detectable IOP changes were observed with the occluded tube outside the eye but within the goggle’s chamber, as shown in Table 3.

**Table 2.**
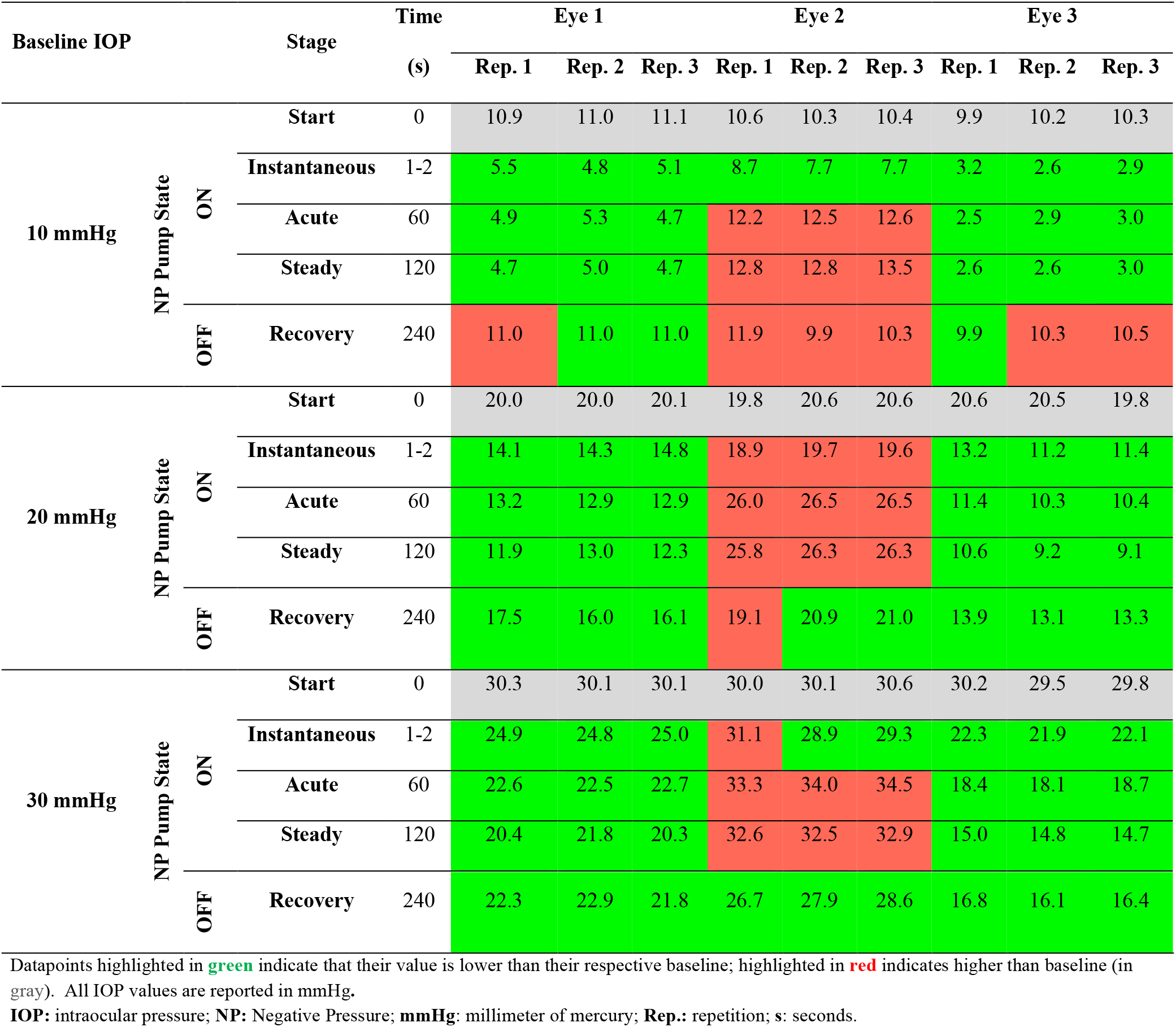
IOP measurements as measured at five time points (0s, 1s, 60s, 120s, 240s) and three baseline IOP (10, 20, 30mmHg) in the three tested eyes in living conditions. Negative pressure was set constant at -20mmHg. The NP pump was ON for the first 120 seconds, then it was turned OFF for the 120 to 240 seconds interval

**Table 3.** Assessment of the effect of negative pressure on the in-line tubing on Eye1

| Stage | Time (s) | Baseline IOP (mmHg) |  |  |
| --- | --- | --- | --- | --- |
|  |  | 10 | 20 | 30 |
| Start | 0 | 10.1 | 20.0 | 30.1 |
| Acute | 60 | 10.1 | 20.0 | 30.1 |
| Steady | 120 | 10.1 | 19.9 | 29.9 |

**NP:** Negative Pressure; **s:** seconds. **IOP:** intraocular pressure; **mmHg:** millimeter of mercury

### Summary statistics

Estimates and p-values for the difference in IOP at the varying stages of negative pressure and baseline IOP are shown in Table 4. The effect of negative pressure on IOP against their baseline was inconsistent across the tested eyes (p>0.05); reduction of IOP was only significant when negative pressure was applied with a baseline IOP of 30mmHg following 120s of exposure (steady state, p<0.001) and remained significantly lower with recovery (p<0.001). The IOP reduction during recovering was also significant at a baseline IOP of 20mmHg (p=0.00113).

**Table 4.**
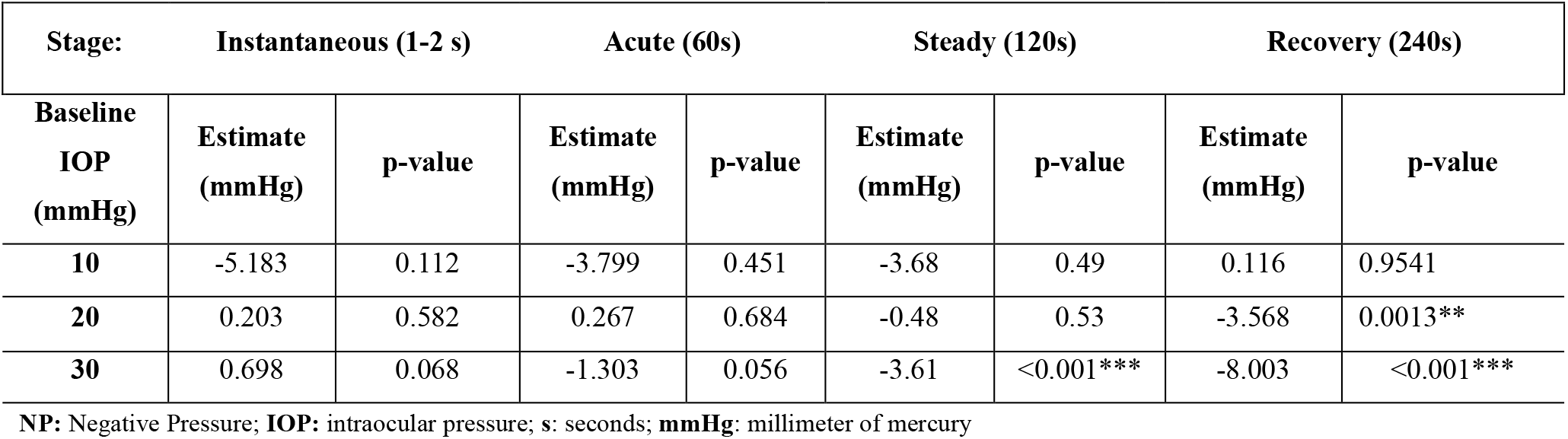
Estimates by linear mixed effect models on the effect of negative pressure on the differential change of IOP from baseline

| Stage: | Instantaneous (1-2 s) |  | Acute (60s) |  | Steady (120s) |  | Recovery (240s) |  |
| --- | --- | --- | --- | --- | --- | --- | --- | --- |
| Baseline IOP (mmHg) | Estimate (mmHg) | p-value | Estimate (mmHg) | p-value | Estimate (mmHg) | p-value | Estimate (mmHg) | p-value |
| 10 | -5.183 | 0.112 | -3.799 | 0.451 | -3.68 | 0.49 | 0.116 | 0.9541 |
| 20 | 0.203 | 0.582 | 0.267 | 0.684 | -0.48 | 0.53 | -3.568 | 0.0013** |
| 30 | 0.698 | 0.068 | -1.303 | 0.056 | -3.61 | <0.001*** | -8.003 | <0.001*** |

NP: Negative Pressure; IOP: intraocular pressure; s: seconds; mmHg: millimeter of mercury

## Discussion

This study is the first to evaluate changes in manometrically-measured intracameral IOP due to negative pressure applied to the ocular surface in living conditions. Results of the study showed a consistent drop in IOP following exposure to negative pressure in two eyes, and an increase in one of the three eyes tested. Upon removal of negative pressure, IOP tended to return to baseline in most of the testing conditions.

The etiology of these conflicting results in IOP lowering by negative pressure is not understood. If the negative pressure is assumed to be distributed around most of the globe, this would induce stretch of the corneoscleral shell with consequential increase of globe volume, which would ultimately result in an acute drop in IOP, as previously predicted in computational models^16^. However, if negative pressure is mostly distributed anteriorly, this negative pressure would primarily induce a protrusion of the corneoscleral shell possibly resulting in a reduction of globe volume, which would ultimately result in an acute increase in IOP. Lastly, unlike the other subjects, the donor with elevated IOP following exposure to negative pressure was morbidly obese (BMI = 52). It is possible that negative pressure distribution around the globe is affected by the amount and location of orbital fat, contributing to the aforementioned reduction of overall globe volume with consequential increase in IOP in eyes. Several studies have examined the impact of atmospheric pressure on IOP with conflicting results. Van de Veire and colleagues^14^ evaluated the impact of changing atmospheric pressure on IOP measured with Perkins handheld applanation tonometer in 27 healthy subjects using a hyperbaric chamber at normal atmospheric pressure (1 Bar) and following elevation of atmospheric pressure increase to 2 Bar and found that there was a slight but significant reduction in IOP [mean reduction: 1.1 mmHg (OD), p=0.024 and 1.4 mmHg (OS), p=0.0006). The reduction in measured IOP remained constant throughout the measurement period of 40 minutes.

Albis-Donado et al.^15^ more recently evaluated the changes in IOP with exposure to increase atmospheric pressure within a hyperbaric chamber with both Goldman applanation tonometry and dynamic contour tonometry. Their study found that with increasing atmospheric pressure, IOP was reduced. However, none of these studies evaluated the absolute IOP, the pressure in the eye relative to vacuum. This is problematic since the elevation of atmospheric pressure itself will directly reduce the IOP as measured across the cornea by applanation tonometry, as transcorneal measured IOP equals the absolute IOP minus atmospheric pressure. This could explain the paradoxical reduction in IOP at increasing atmospheric pressure.

Application of the negative pressure goggles has been proposed as a novel non-medical and non-invasive treatment for glaucoma and recent computational modeling has predicted that IOP will reduce with the application of negative pressure goggles initially due to scleral expansion and longer term with increasing episcleral outflow.^16^ Another recent computational study reported a reduction in IOP with exposure to negative pressure^24^, which is consistent with the expected mechanical response of a generic closed vessel under pressure expose to reduce external pressure. Results of numerical models based on simplified and generic ocular morphology and mechanical properties would of course omit all the eye-specific and orbital characteristics that may mediate the response of IOP to varying extra orbital pressure, as observed by our study.

The short and prolonged impact of negative pressure has been evaluated in living patients using a modified version of these goggles to allow for pneumotonometry through a silicon membrane.^20, 21, 25^ These studies showed similar changes in IOP to our current study in two of our non-obese subjects. However, none of the subjects in these prior clinical studies experienced an elevation in IOP. Measurements of BMI and weight were not included in these studies.

Kamalipour et al., recently evaluated the impact of the negative pressure goggles on retinal and ONH perfusion.^17^ This study examined 24 subjects with open angle glaucoma, using OCTA to determine the impact of negative pressure on measurements of perfused capillary density (PCD) and retinal nerve fiber layer (RNFL) thickness. Prior studies examining the relationship between IOP and retinal perfusion have demonstrated increased retinal blood flow following sustained lowering of IOP using laser doppler flowmetry^26^ and dynamic alteration in blood flow during vitrectomy using and intraocular transducer ^27^, with increased flow associated with decreased IOP. OCTA has previously been used to evaluate the chronic impact of surgical IOP lowering on capillary density in several studies, which show increased capillary density with lowering of IOP ^28-31^. Thus, the authors sought to utilize OCTA to evaluate the tissues effect of negative pressure as a surrogate for measuring absolute pressure. While the authors did not find a change in RNFL thickness, they reported that, overall, there was an increase in perfused capillary density with increasing negative pressure for the group, which decreased as negative pressure was removed. While these results seem to suggest that IOP was reduced with negative pressure in their study subjects, they are not discordant with our results. In the prior clinical study, the mean changes in PCD seen at peak negative pressure was 2.3%, with 95% confidence intervals of 0.5% to 4%. While the range was not provided, given this distribution, it is probable that negative pressure reduced capillary density in a minority of observations. Nevertheless, compression of the vasculature tissue following an acute increase in IOP (as shown by our group^32^) could mechanistically induce an increase in detected PCD by OCTA because of an increase in blood velocity (a parameter of influence in OCTA) and/or induce an overall reduction of vasculature tissue volume, which would displace peripheral blood volume. Lastly, body mass index, a potential mediator for IOP change and negative pressure, was not reported for the study subjects, thus the study may not have included morbidly obese individuals with high orbital fat content.

Ultimately, retrobulbar orbital tissue may play a role in modulating the amount and extent of negative pressure reaching the posterior segment of the eye following application of negative pressure to the anterior segment of the eye. The retrobulbar space is filled by an inhomogeneous combination of fluid, fat, muscles, and tissue. As such, it is erroneous to describe its mechanical state by simply measuring compressive pressure within the tissue, as done by a recent study measuring the effect of negative pressure on retrobulbar pressure in human cadaver eyes.^33^ Compressive pressure is only one of the three principal components of the strain tensor defining the mechanical state of a deformable tissue; it is descriptive of a mechanical state only when the tissue is exposed to purely hydrostatic forces, which is not the case for retrobulbar tissue.

Overall, our results support the concept that negative pressure can lower IOP in most subjects, as there was a relatively consistent lowering of IOP in the remaining non-obese donors that is consistent with the pressure reductions seen with modified excursion goggles and in computational models. However, additional studies would be required to determine the relationship between BMI, orbital fat, and the ocular response to negative pressure. Ideally, clinical studies evaluating this approach as a treatment should consider including obese subjects to determine if there is paradoxical elevation of IOP in these subjects.

The current study has several limitations. First, given the constraints of the setting in these donors, more prolonged evaluation of negative pressure was not possible. Also, indwelling IOP sensors were not used. However, there was no pressure change seen when the tube was occluded, thus change induced from negative pressure on the tubing within the negative pressure compartment is unlikely to have had a significant impact on the change in absolute IOP. Second, given the limited availability of research consented organ donors, it is difficult to perform testing on and extensive number of donors. Third, magnitude of negative pressure was nominally set by the MPD pressure pump; actual magnitude of the effective negative pressure within the goggles was not independently measured. Lastly, we were not able to perform imaging of the ONH and retina during the application of negative pressure due to time constrains. Seminal studies are ongoing examining the impact of longer-term exposure to negative pressure on absolute IOP and globe expansion using indwelling piezoelectric sensors and OCT in living condition and ex vivo inflation testing of the corneoscleral shell to assess the effect of negative pressure on the ONH and retina.

## Conclusions

This study has directly examined the impact of negative pressure applied to the ocular surface on absolute IOP for the first time in the living human eye. While there was consistent lowering of absolute IOP with application of simulated negative pressure in two of the tested eyes, it is concerning that the eye from a donor with high BMI had significantly elevated absolute IOP following exposure to negative pressure. We hypothesize that orbital fat may modulate the interaction between negative pressure and globe expansion by either preventing uniform distribution of negative pressure around the globe (necessary for inducing globe expansion - hence IOP reduction) or by inducing compressive pressure on portions of the globe counterbalancing its expansion. Regardless of the etiology, further studies are needed that include patients with elevated BMI before widespread clinical application of this therapeutic approach is applied in glaucoma patients.

## Data Availability

All data produced in the present study are available upon reasonable request to the authors

## Abbreviations

(IOP): Intraocular Pressure;
(MPD): Multi-Pressure Dial;
(BDOD): Brain-Dead Organ Donors;
(BSS): Balanced Salt Solution;
(OCT): Optical Coherence Tomography;
(OCTA): Optical Coherence Tomography Angiography;
(ONH): Optic Nerve Head;
(CSFP): Cerebrospinal Fluid Pressure;
(RNFL): Retinal Nerve Fiber Layer;
(PCD): Perfused Capillary Density;

## Support

*National Eye Institute*, R01 EY028284: Financial support

*Heidelberg Engineering, GmbH*: Hardware support

*Topcon Healthcare System, Inc*.: Financial and hardware support

*Research to Prevent Blindness*: Unrestricted grant

*Eyesight Foundation of Alabama*: Unrestricted grant

## Royalties or licenses

*Equinox Ophthalmic, Inc*.: Licensing fees for part of the dataset

## Patents

*Colocalized detection of retinal perfusion and optic nerve head deformations*, WO2019178185A1

*Retinal vascular stress test for diagnosis of vision-impairing diseases*, WO2021158903A1

## Notes

### Competing Interest Statement

Heidelberg Engineering, GmbH: Hardware support
Topcon Medical Systems, Inc.: Financial and hardware support
Equinox Ophthalmic, Inc.: Licensing fees for part of the published dataset
Patent: Colocalized detection of retinal perfusion and optic nerve head deformations, WO2019178185A1
Patent: Retinal vascular stress test for diagnosis of vision-impairing diseases, WO2021158903A1

### Funding Statement

National Eye Institute, R01 EY028284
Research to Prevent Blindness (unrestricted grant)
EyeSight Foundation of Alabama (unrestricted grant)

### Author Declarations

All components of this study adhered to the Declaration of Helsinki and were approved by the UAB Institutional Review Board and Legacy of Hope Research Review Board.

### Summary of Updates

The revised version slightly modifies Figure 2 and Table 2 for more immediate interpretation. Two labels in Tables 3 and 4 are then edited to align them with the edits of Fig. 2 and Table2. Line spacing in the Discussion was also discordant with the other sections. All terms are now capitalized in Abbreviations.

